# Covid-19 does not look like what you are looking for: Clustering symptoms by nation and multi-morbidities reveal substantial differences to the classical symptom triad

**DOI:** 10.1101/2021.04.02.21254818

**Authors:** Balasundaram Kadirvelu, Gabriel Burcea, Jennifer K Quint, Ceire E Costelloe, A. Aldo Faisal

## Abstract

COVID-19 is by convention characterised by a triad of symptoms: cough, fever and loss of taste/smell. The aim of this study was to examine clustering of COVID-19 symptoms based on underlying chronic disease and geographical location. Using a large global symptom survey of 78,299 responders in 190 different countries, we examined symptom profiles in relation to geolocation (grouped by country) and underlying chronic disease (single, co- or multi-morbidities) associated with a positive COVID-19 test result using statistical and machine learning methods to group populations by underlying disease, countries, and symptoms. Taking the responses of 7980 responders with a COVID-19 positive test in the top 5 contributing countries, we find that the most frequently reported symptoms differ across the globe: For example, fatigue 4108(51.5%), headache 3640(45.6%) and loss of smell and taste 3563(44.6%) are the most reported symptoms globally. However, symptom patterns differ by continent; India reported a significantly lower proportion of headache (22.8% vs 45.6%, p<0.05) and itchy eyes (7.0% vs. 15.3%, p<0.05) than other countries, as does Pakistan (33.6% vs 45.6%, p<0.05 and 8.6% vs 15.3%, p<0.05). Mexico and Brazil report significantly less of these symptoms. As with geographic location, we find people differed in their reported symptoms, if they suffered from specific underlying diseases. For example, COVID-19 positive responders with asthma or other lung disease were more likely to report shortness of breath as a symptom, compared with COVID-19 positive responders who had no underlying disease (25.3% vs. 13.7%, p<0.05, and 24.2 vs.13.7%, p<0.05). Responders with no underlying chronic diseases were more likely to report loss of smell and tastes as a symptom (46%), compared with the responders with type 1 diabetes (21.3%), Type 2 diabetes (33.5%) lung disease (29.3%), or hypertension (37.8%). Global symptom ranking differs markedly from the well-known and commonly described symptoms for COVID-19, which are based on a few localised studies. None of the five countries studied in depth recorded cough or temperature as the most common symptoms. The most common symptoms reported were fatigue and loss of smell and taste. Amongst responders from Brazil cough was the second most frequently reported symptom, after fatigue. Moreover, we find that across countries and based on underlying chronic diseases, there are significant differences in symptom profiles at presentation, that cannot be fully explained by the different chronic disease profiles of these countries, and may be caused by differences in climate, environment and ethnicities. These factors uncovered by our global comorbidity survey of COVID-19 positive tested people may contribute to the apparent large asymptotic COVID-19 spread and put patients with underlying disease systematically more at risk.

**Executive Summary:** *Evidence before this work:* An early meta-analysis of epidemiological variation in COVID-19 inside and outside China studied patient characteristics including, gender, age, fatality rate, and symptoms of fever, cough, shortness of breath and diarrhoea in COVID-19 patients. They found that important symptom differences existed in patients in China compared to other countries and recommended that clinical symptoms of COVID-19 should not be generalized to fever, shortness of breath and cough only, but other symptoms such as diarrhoea are also shown to be prevalent in patients with COVID-19.

*Added value of this work:* *W*e find that across countries and based on underlying chronic diseases, there are significant differences in symptom profiles at presentation, that cannot be fully explained by the different chronic disease profiles of these countries, and may be caused by differences in climate, environment and ethnicities.

*Implications of the evidence:* These factors, uncovered by our global comorbidity survey of COVID-19 positive tested people may contribute to the apparent large asymptotic COVID-19 spread and put patients with underlying disease systematically more at risk.

## INTRODUCTION

Since the start of the COVID-19 pandemic, most testing has been triggered by a classical triad of symptoms, which were first observed in COVID-19 patients who were hospitalised. However, grouping patients based on clinical characteristics is crucial for clinical practice, to allow selection of diagnostic tests and to predict prognosis. Web-based symptom checkers have become popular in the context of the novel COVID-19 pandemic, as access to physicians is reduced, concern in the population is high, and large amounts of misinformation are circulating social media. (1) On COVID-19 symptom checker web pages, users are asked a series of COVID-19–specific questions; upon completion, an association between the answers and COVID-19 is given alongside in some instances, behavioural recommendations, such as self-isolation. In this context, COVID-19 symptom checkers are valuable tools for pre-assessment and screening.

However, with COVID-19, important biological differences are likely to exist between patient subgroups, as is seen in other forms of critical illness. This has been demonstrated where highly significant sub-group effects were observed in the first drug trial to demonstrate an improvement in mortality, dexamethasone.(2) However, in COVID-19, important biological differences are likely to exist between patient subgroups and contexts. People in different locations/cultures may perceive symptoms differently, and underlying diseases may mask/alter certain “typical” covid symptoms. Additionally, sub-groups of the population are at higher risk of both developing COVID-19 and experiencing more severe infection, people aged 60 years and older; those living in long-term care facilities; and people with underlying health conditions, such as hypertension, diabetes, cardiovascular disease, chronic respiratory disease and weakened immune systems. On a global level, as new variants of COVID-19 arise in different parts of the world, it is hypothesised that symptom profiles may differ according to variant, and indeed geographical location. (3)

It is important to characterise the symptoms of severe COVID-19 and to identify clinical sub-groups. This will speed up diagnosis, enable more precise prediction of outcomes, and target treatment. The aim of this study was to examine clustering of COVID-19 symptoms based on underlying chronic disease and geographical location.

## METHODS

### Study setting and participants

In April 2020, the company Your.MD launched a web-based COVID-19 Symptom Mapper (4) in partnership with Imperial College London’s Global Covid Observatory to better understand how the disease is affecting communities worldwide. The symptom mapper allows participants to complete a survey on COVID-19 symptoms, any outcome of COVID-19 testing and recording underlying conditions as well as pre-specified risk factors for COVID-19. All survey questions are laid out in Supplementary Table S1. As of 22nd September 2020, 175,572 people around the world used the mapper to record their symptoms.

### Data Curation

The study made use of the Your.MD symptom mapper data (4) collected globally between 09/04/2020 and 22/09/2020. Of the 175,571 total responders in the collected data, only 78,299 responders whose age was in the range 0-100 years and chose tested positive(n=7980), tested negative (n=5620), or showing symptoms (n= 64,699) for the reason for participating in the survey were considered for further analysis. All symptoms were mapped to a binary categorical variable for analysis. “Sneezing” was excluded from the list of symptoms presented as none of the tested positive individuals experienced it.

### Statistical analysis

Characteristics of the responding cohort were described and visualised using histograms and heatmaps. Frequency counts of symptoms were described and plotted, stratified by age and underlying chronic disease. Radar plots were used for visual comparison of symptom profiles and visualise significant differences between profiles. A comparative analysis was conducted to examine the association between symptoms, and groups of symptoms with geographical location of respondents and underlying chronic diseases reported. We compared the proportions using two-sided χ2 test at a significance level of 5% with Bonferroni correction applied.

Multivariable logistic regression was used including *a priori* specified confounding variables. For the multivariate analysis, age was converted to a categorical variable (>60 years). The gender variable was coded as 0 for male and 1 for female. To cluster the symptoms, we used agglomerative hierarchical clustering of the symptoms with the Jaccard dissimilarity measure as the distance and unweighted average distance as the linkage criteria. The Jaccard dissimilarity measure is defined as one minus the Jaccard coefficient, which is the percentage of nonzero symptoms that differ. (5) For clustering, we excluded symptoms experienced by less than 5% of the individuals in each group to avoid chaining.(6) All statistical analyses were performed using custom programs in the MATLAB R2019b (MathWorks) environment.

### Ethics

The Covid-10 Symptom Mapper data was provided to us by Your.MD free of charge and obligations with freedom to publish any results. The data is provided on request for free from Your.MD. On the Your.MD website all participants provided informed consent at the start of the online questionnaire for their data to be used for research purposes, and had to agree to the corresponding Your.MD privacy and data usage policies [Your.MD]

## RESULTS

Between 09/04/2020 and 22/09/2020 175,566 individuals from 190 different countries responded to the COVID-19 YOUR.MD questionnaire. The countries with the highest proportion of respondents (Figure 1) were India (39590, 22.5%), Mexico (29644, 16.8%), Pakistan (16820, 9.5 %), Philippines (1590, 9.0 %), United Kingdom (10044, 5.7 %), and Brazil (9796, 5.5%), accounting for 63.8 % of the total number of responders. The top-5 countries with the highest percentage of individuals who tested positive were Mexico (2199, 27.6%), Brazil (1366, 17.1%), Pakistan (753, 9.4%), India (714, 8.9%) and United Kingdom (420, 5.3%) and the detailed descriptive statistics for responders from Mexico, Brazil, Pakistan, India and United Kingdom are presented in Supplementary Table S2.

**Figure 1:**
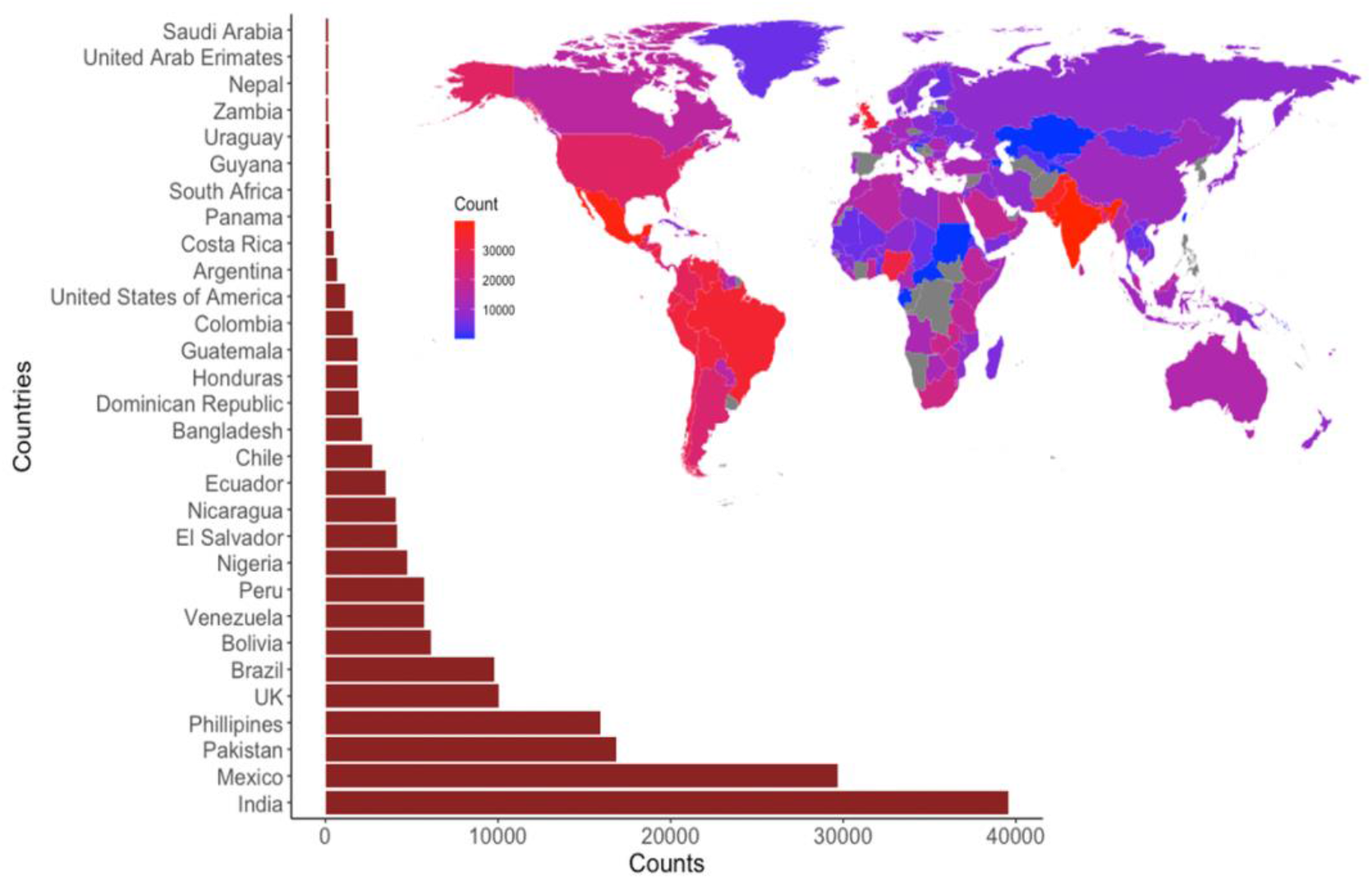
Global response to COVID-19 symptom survey; counts by country.

Table 1 describes the characteristics of responders who reported: a COVID-19 positive test; a COVID-19 negative test result, or that they were showing COVID-19 symptoms. Data from responders with unknown COVID-19 status (e.g. ‘self-isolating with no symptoms’, ‘curious’, ‘live with someone with corona virus’), were excluded from the analysis, resulting in 78,299 respondents included in the analysis. Of these responders 7980 (10.2 %) tested positive, 5620 (7.2%) tested negative, and 64699 (82.6 %) reported symptoms but had not been tested. In the overall cohort mean age was 35.9(11.6), responders who tested positive and negative for COVID-19 were older than responders who reported symptoms but had not been tested. A higher proportion of women completed the survey (59.3%). There was a higher proportion of health care workers who had been tested and reported a COVID-19 positive or COVID-19 negative test compared with those responders who were showing symptoms (18.1% vs 7.7%). Responders had been experiencing COVID-19 symptoms for a median of 4 days, with responders who were COVID-19 positive reporting a longer duration of symptoms (median, 7 days) compared with responders who had symptoms but had not been tested (median, 4 days).

**Table 1:** Characteristics of responders to Your.MD symptom questionnaire, categorised as tested positive, showing symptoms (but not tested), or tested negative

| SARS-Covid-19 status |  |  |  |  |
| --- | --- | --- | --- | --- |
|  | All responders | Tested Positive | Untested Symptomatic | Tested Negative |
| **Count** | 78299 | 7980 | 64699 | 5620 |
| **Age mean(SD)** | 35.9(11.6) | 38.9(11.5) | 35.3(11.6) | 37.7(11.8) |
| Females (%) | 46451(59.3) | 4993(62.6) | 38183(59.0) | 3275(58.3) |
| Pregnant (%) | 1137(1.5) | 137(1.7) | 915(1.4) | 85(1.5) |
| Care home worker (n/%) | 6925(8.8) | 725(9.1) | 5621(8.7) | 579(10.3) |
| Health care worker (n/%) | 7354(9.4) | 1438(18.0) | 4896(7.6) | 1020(18.1) |
| Number of days of symptoms (mean/std) | 6.3(7.6) | 7.9(7.3) | 6.1(7.6) | 6.3(8.0) |
| Number of days of symptoms (median/IQR) | 4.0(5.0) | 7.0(7.0) | 4.0(5.0) | 4.0(6.0) |
| **Brazil** |  |  |  |  |
| **Count** | 7257 | 1366 | 5089 | 802 |
| **Age mean (SD)** | 38.4(11.3) | 40.6(10.9) | 37.5(11.3) | 40.0(11.3) |
| Females (%) | 5909(81.4) | 1133(82.9) | 4127(81.1) | 649(80.9) |
| Pregnant (%) | 92(1.3) | 17(1.2) | 67(1.3) | 8(1.0) |
| Care home worker (n/%) | 265(3.7) | 51(3.7) | 175(3.4) | 39(4.9) |
| Health care worker (n/%) | 868(12.0) | 241(17.6) | 476(9.4) | 151(18.8) |
| Number of days of symptoms (mean/std) | 5.3(5.6) | 8.4(7.0) | 4.3(4.6) | 6.3(6.9) |
| Number of days of symptoms (median/IQR) | 4.0(5.0) | 8.0(7.0) | 3.0(4.0) | 5.0(6.0) |
| **India** |  |  |  |  |
| **Count** | 8967 | 714 | 7552 | 701 |
| **Age mean(SD)** | 34.4(11.5) | 38.1(12.4) | 34.1(11.4) | 34.4(11.7) |
| Females (%) | 3415(38.1) | 282(39.5) | 2901(38.4) | 232(33.1) |
| Pregnant (%) | 150(1.7) | 21(2.9) | 115(1.5) | 14(2.0) |
| Care home worker (n/%) | 1078(12.0) | 121(16.9) | 838(11.1) | 119(17.0) |
| Health care worker (n/%) | 634(7.1) | 84(11.8) | 448(5.9) | 102(14.6) |
| Number of days of symptoms (mean/std) | 7.3(9.7) | 6.7(7.4) | 7.4(9.8) | 7.5(10.9) |
| Number of days of symptoms (median/IQR) | 4.0(6.0) | 5.0(8.0) | 4.0(5.0) | 3.0(7.0) |
| **Mexico** |  |  |  |  |
| **Count** | 20616 | 2199 | 17432 | 985 |
| **Age mean(SD)** | 35.9(10.8) | 38.5(10.6) | 35.5(10.7) | 37.9(10.8) |
| Females (%) | 13138(63.7) | 1385(63.0) | 11156(64.0) | 597(60.6) |
| Pregnant (%) | 244(1.2) | 26(1.2) | 211(1.2) | 7(0.7) |
| Care home worker (n/%) | 1196(5.8) | 132(6.0) | 990(5.7) | 74(7.5) |
| Health care worker (n/%) | 1706(8.3) | 357(16.2) | 1194(6.8) | 155(15.7) |
| Number of days of symptoms (mean/std) | 5.4(5.6) | 8.0(6.7) | 5.1(5.4) | 5.2(5.5) |
| Number of days of symptoms (median/IQR) | 4.0(5.0) | 7.0(7.0) | 4.0(5.0) | 4.0(5.0) |
| **Pakistan** |  |  |  |  |
| **Count** | 7451 | 753 | 6286 | 412 |
| **Age mean (SD)** | 31.9(10.8) | 36.1(12.2) | 31.3(10.6) | 33.0(10.5) |
| Females (%) | 3391(45.5) | 283(37.6) | 2934(46.7) | 174(42.2) |
| Pregnant (%) | 230(3.1) | 29(3.9) | 188(3.0) | 13(3.2) |
| Care home worker (n/%) | 1447(19.4) | 119(15.8) | 1244(19.8) | 84(20.4) |
| Health care worker (n/%) | 851(11.4) | 160(21.2) | 588(9.4) | 103(25.0) |
| Number of days of symptoms (mean/std) | 5.0(6.1) | 6.4(5.7) | 4.8(6.2) | 4.7(5.9) |
| Number of days of symptoms (median/IQR) | 3.0(4.0) | 5.0(6.0) | 3.0(4.0) | 3.0(5.0) |
| **United Kingdom** |  |  |  |  |
| **Count** | 2977 | 420 | 2379 | 178 |
| **Age mean (SD )** | 47.3(13.1) | 46.9(9.8) | 47.3(13.7) | 47.0(12.4) |
| Females (%) | 2290(76.9) | 342(81.4) | 1812(76.2) | 136(76.4) |
| Pregnant (%) | 26(0.9) | 1(0.2) | 24(1.0) | 1(0.6) |
| Care home worker (n/%) | 297(10.0) | 108(25.7) | 166(7.0) | 23(12.9) |
| Health care worker (n/%) | 567(19.0) | 192(45.7) | 310(13.0) | 65(36.5) |
| Number of days of symptoms (mean/std) | 10.2(11.1) | 10.1(8.4) | 10.1(11.3) | 12.1(13.2) |
| Number of days of symptoms (median/IQR) | 6.0(11.0) | 8.0(8.0) | 5.0(12.0) | 6.0(17.0) |

Table 2 describes symptoms reported by COVID-19 positive responders. Responders who tested positive were more likely to report joint pain (7% vs 5.2%), loss of appetite (13.1% vs 8%) and loss of smell and taste (44.6% vs 32.4%) than responders who had tested negative or had symptoms but had not been tested. Fewer responders who had tested positive for COVID-19 reported sore throat as a symptom (30.2% vs 42.8%) compared with the overall population of responders. Of responders who were COVID-19 positive, 30.6 % have one underlying chronic disease, 6.11 % reported 2 underlying chronic diseases, and 1.52 % reported 3 or more. The presence of underlying chronic disease was similar in the COVID-19 positive responders compared with the overall responder population. The detailed descriptive statistics for comorbidities are listed in Supplementary Table S3.

**Table 2:**
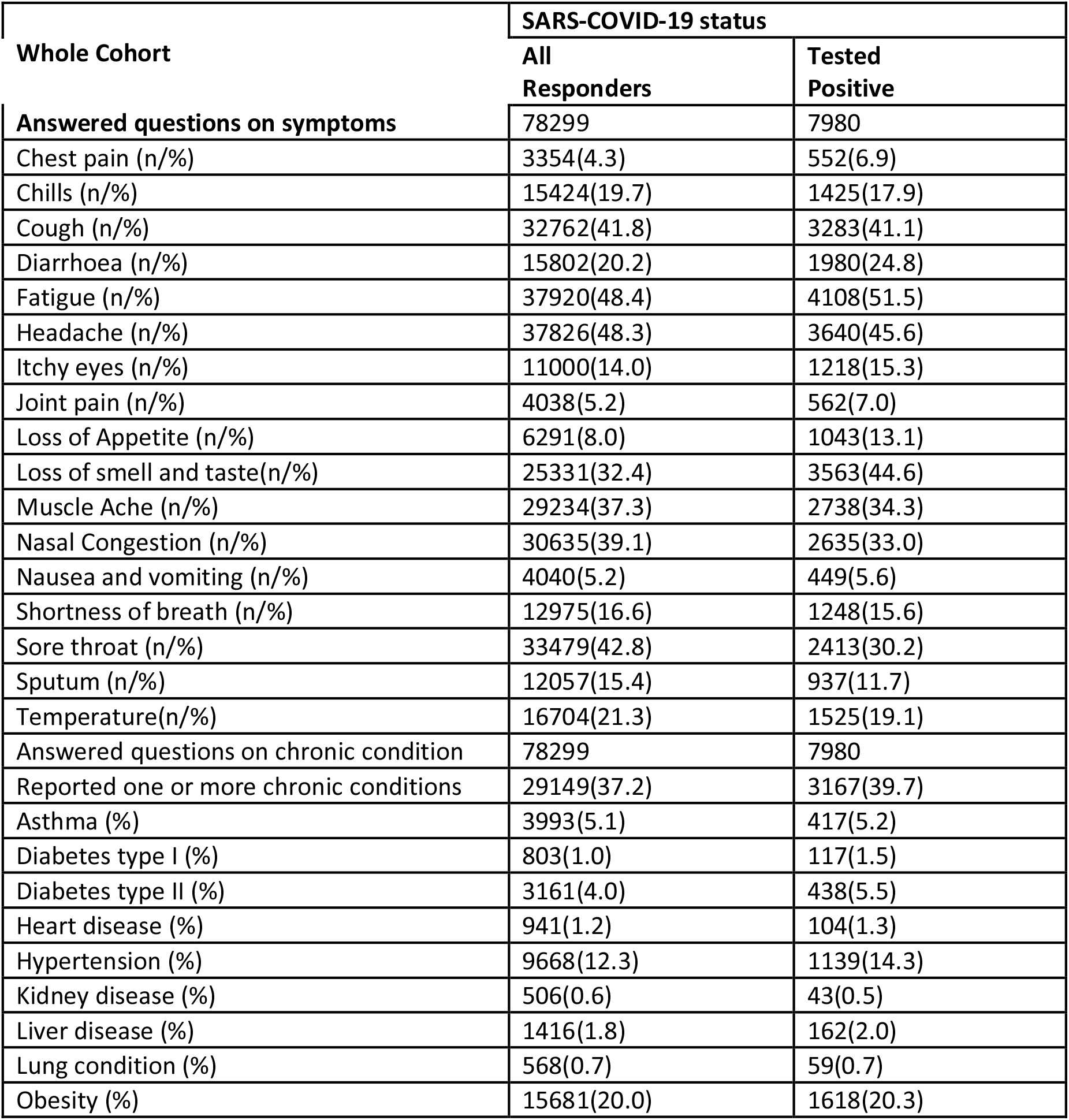
Symptoms and comorbidities reported by Your.MD questionnaire responders.

**Table 3:** Comparison of most reported symptoms (normally >33%) in our global survey and major and minor symptoms as stated by selected national or international publications.

| Country | This global survey (only top 3 prevalence, *indicates significant difference from other countries) | Official main/most common symptoms | Official additional/Less common symptoms | Ref |
| --- | --- | --- | --- | --- |
| Brazil | •headache*,<br>•loss of appetite*,<br>•fatigue<br>•cough | •fever,<br>•tiredness,<br>•dry cough | •pain,<br>•nasal congestion,<br>•headache,<br>•conjunctivitis,<br>•sore throat,<br>•diarrhea,<br>•loss of taste or smell,<br>•rash on the skin or discoloration of the fingers or toes | (7) |
| India | •fever (temperature)<br>•cough<br>•fatigue* | •fever,<br>•tiredness,<br>•dry cough. | •aches and pains,<br>•nasal congestion,<br>•runny nose, | (8) |
|  | •headache*,<br>•loss of smell or taste* |  | •sore throat<br>•diarrhoea |  |
| Pakistan | •chills*<br>•headache*,<br>•fatigue<br>•loss of smell or taste<br>•sore throat | • Respiratory symptoms (cough and shortness of breath)<br>• Fever<br>• Muscle pain<br>• Headache<br>• Sore throat<br>• New loss of taste or smell<br>• Severe fatigue |  | (9) |
| WHO | [reporting global statistics as proxy]<br>•fatigue<br>•cough<br>•loss of smell or taste<br>•sore throat<br>•nasal congestion | • Fever<br>• Dry cough<br>• Fatigue | • Loss of taste or smell,<br>• Nasal congestion,<br>• Conjunctivitis (also known as red eyes)<br>• Sore throat,<br>• Headache,<br>• Muscle or joint pain,<br>• Different types of skin rash,<br>• Nausea or vomiting,<br>• Diarrhea,<br>• Chills or dizziness. | (10) |
| Mexico | •cough<br>•fatigue*<br>•headache*<br>•loss of smell and taste*<br>•muscle ache*<br>•sore throat | •headache,<br>•arthralgia or myalgia,<br>•sore throat. | •abdominal pain,<br>•breathing difficulty,<br>•chills,<br>•conjunctivitis,<br>•constant dry cough,<br>•diarrhoea,<br>•fever,<br>•runny nose,<br>•vomiting | (16) |
| UK | •chills*<br>•cough*<br>•fatigue*<br>•nasal congestion*<br>•muscle ache*<br>•shortness of breath* | •new persistent cough;<br>•high temperature,<br>•loss of taste and smell. |  | (17) |

Figure 2 shows the age distribution of responders with and without a chronic disease who were COVID-19 positive, for all countries and the top five responding countries. The second column shows the counts of symptoms for individuals who were COVID-19 positive, for all countries and the top five responding countries. The age profile of the responding population was centred around 20-29 year old age band. Responders from the UK were on average older than other countries. Mexico had the highest prevalence of comorbidities particularly hypertension and obesity compared with other represented countries.

**Figure 2.**
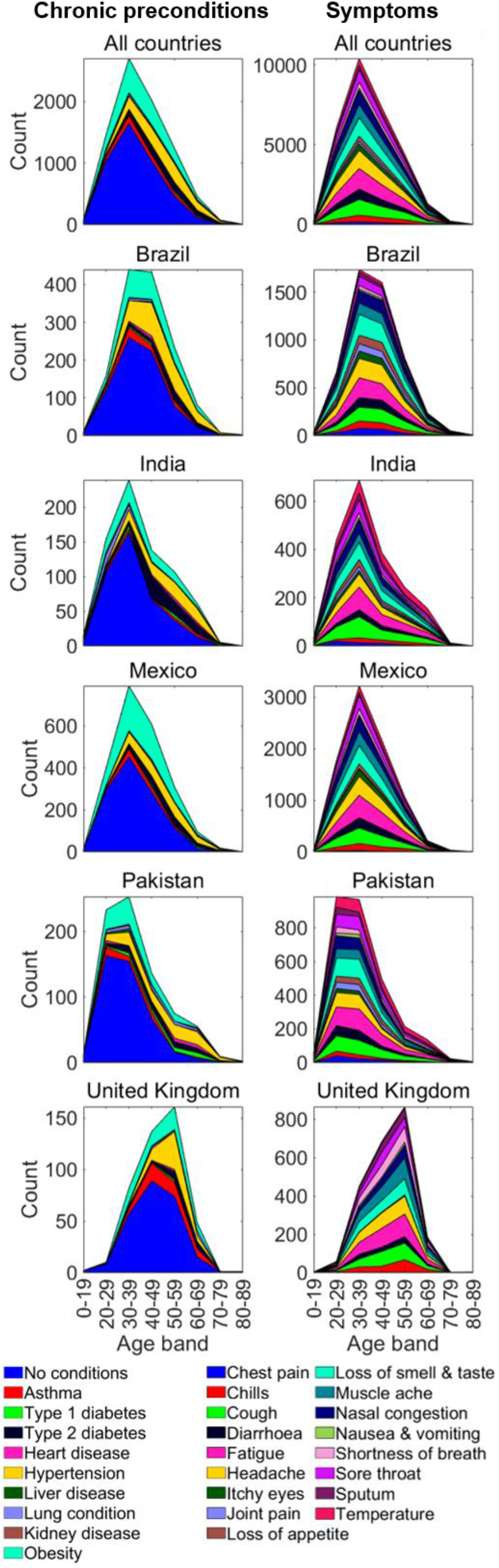
Distribution of symptoms and underlying chronic diseases of COVID-19 tested positive responders from the entire cohort and the top 5 countries.

To further explore the difference in symptom profiles associated with underlying chronic disease and location, a series of radar plots were produced to visualise multivariate data. Figure 3.a shows these symptom profiles amongst COVID-19 positive responders with and without underlying chronic disease.

**Figure 3.**
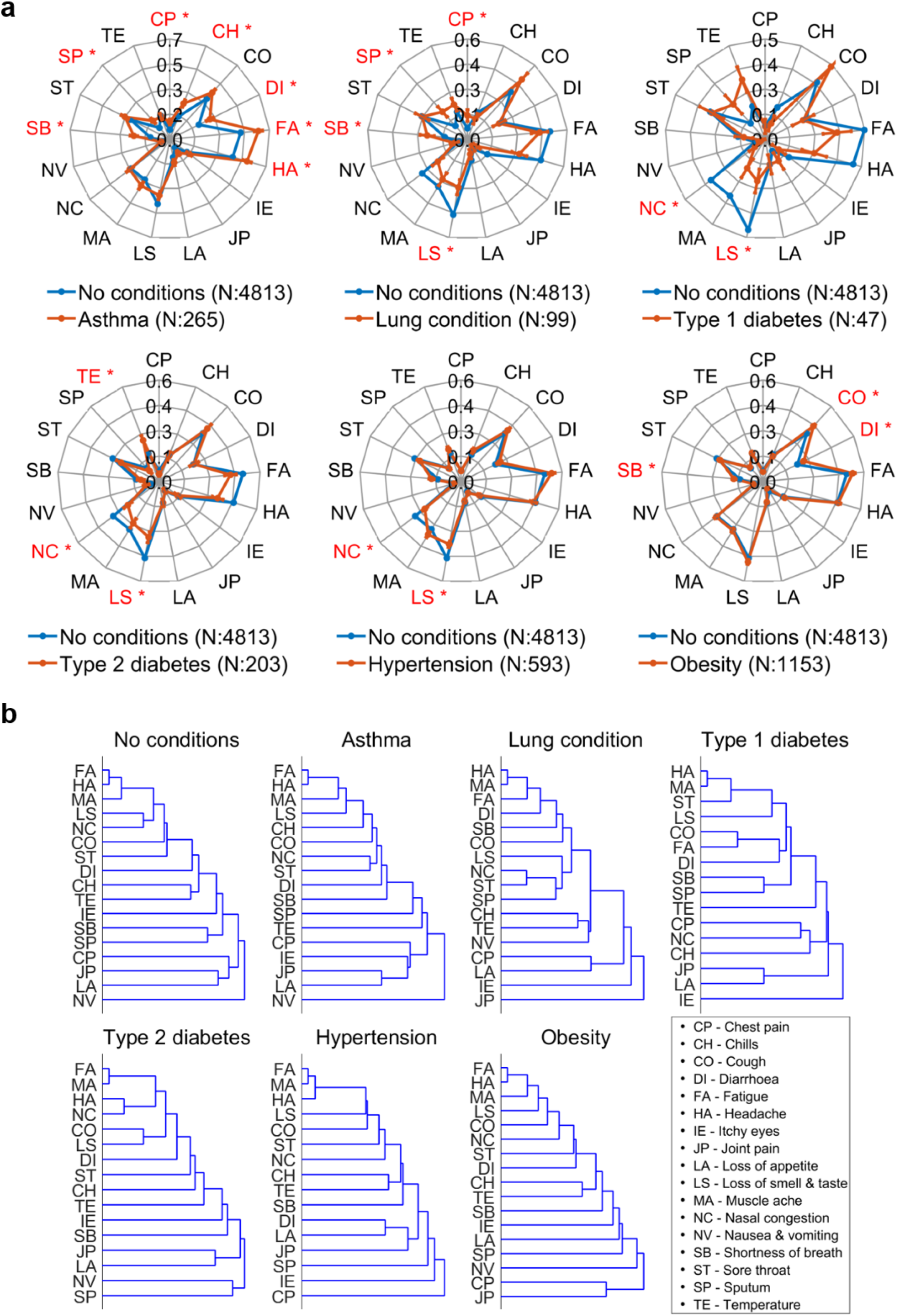
a. Symptom profile difference between no disease group and each chronic disease group. Error bars shown are standard errors and * denotes statistically significant difference (p<0.05, two-sided χ2 test). b. Dendrograms of clustering of the symptoms for different comorbidity groups amongst tested positive responders

A dendrogram in Figure 3.b showing clustering of symptoms across COVID-19 positive responders with and without underlying diseases shows that fatigue, muscle ache and headache were the most commonly reported symptoms. Differences in symptom profile between those with an underlying disease, and those without were evident across all comorbidity groups. For example, COVID-19 positive responders with asthma were more likely to report fatigue, headache, shortness of breath, sputum production, chest pain, chills or diarrhoea, compared with COVID-19 responders who had no underlying disease. COVID-19 positive responders with an underlying lung condition were more likely to report shortness of breath as a symptom, but less likely to report loss of smell and taste compared with COVID-19 positive responders who had no underlying disease. Amongst COVID-19 positive responders with Type 2 Diabetes, a raised temperature was more likely to be reported. Loss of smell and taste was more likely to be reported as a symptom amongst COVID-19 positive responders with no underlying disease compared with responders who had a lung condition, Type 1 diabetes, Type 2 diabetes or hypertension. Symptom profiles of the chronic diseases with three smallest sample sizes (heart, liver and kidney conditions) are presented in Supplementary Figure S1.

Figure 4 shows differences in symptom profile from COVID-19 positive responders across countries. Brazil and Mexico reported a higher number of COVID-19 positive responders with itchy eyes and headache, but fewer reports of sputum or shortness of breath compared with other countries. India and Pakistan reported fewer responders COVID-19 positive with nasal congestion, muscle ache of loss of smell and tastes, compared with other countries.

**Figure 4.**
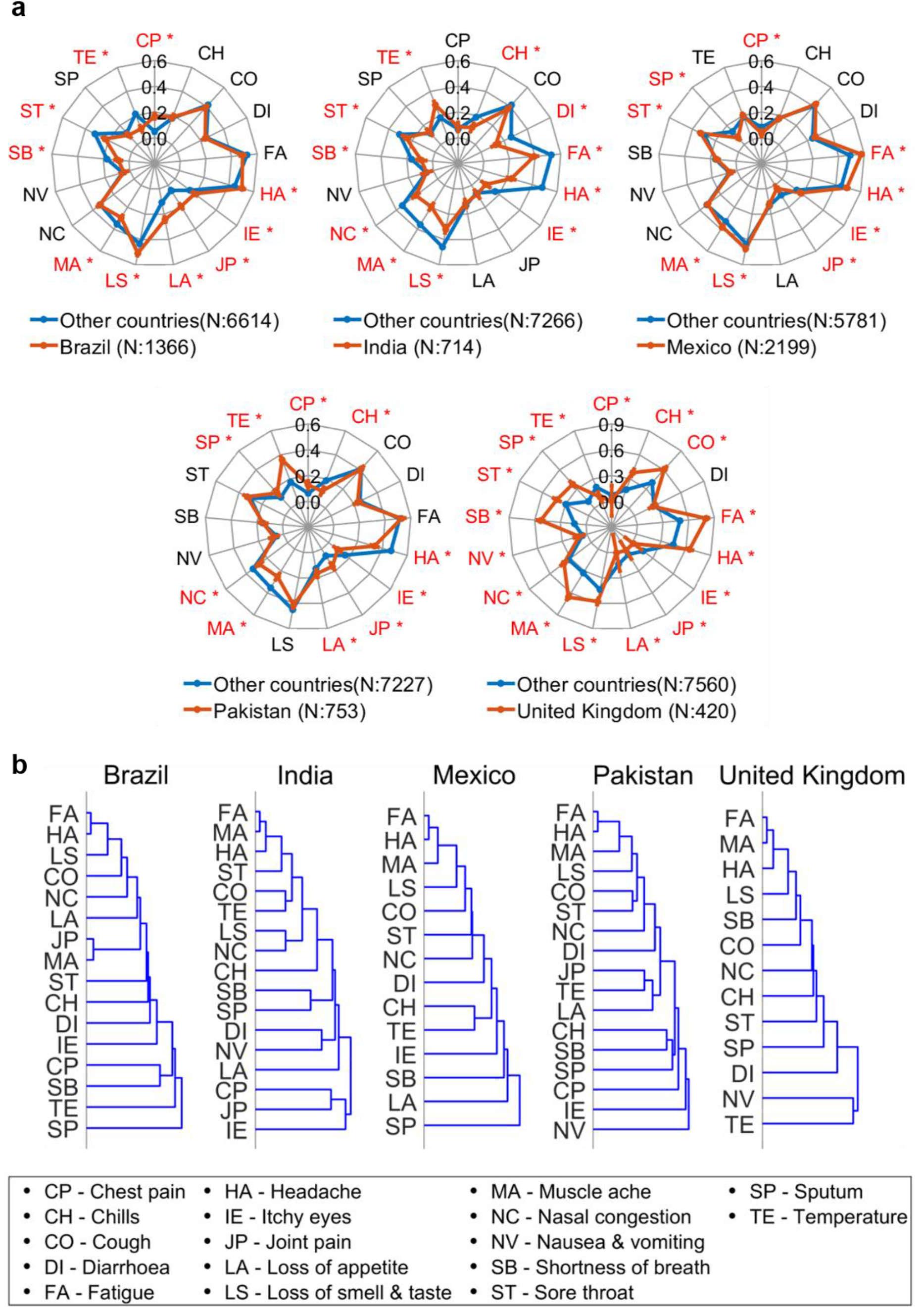
a. Radar plot of the symptom profiles from different countries. Error bars shown are standard errors and * denotes statistically significant difference (p<0.05, two-sided χ2 test). b. Dendrograms of clustering of the symptoms for different countries amongst tested positive responders

A higher number of COVID-19 positive responders in Brazil reported itchy eyes, loss of smell and taste, appetite loss, joint pain, headache and chest pain compared with other countries. A higher number of COVID-19 positive responders in India reported a high temperature as a symptom, compared with other countries. In Mexico, a higher number of COVID-19 positive responders reported muscle ache, itchy eyes, fatigue, sputum and headache compared with other countries. A higher number of COVID-19 positive responders in Pakistan reported a high temperature, joint pain or chest pain compared with other countries. Differences were most marked for COVID-19 positive responders from the UK. A higher number of responders reported loss of smell and taste, muscle ache, shortness of breath, sputum, sore throat, chills, cough, fatigue and headache compared with other countries. This may reflect the differing age range of the UK responder population.

Symptoms profiles for COVID-19 positive patients varied based on and underlying chronic disease and country. A multivariate logistic regression adjusting for age and gender was applied to investigate the association between each of the symptoms and the chronic disease and country of residence and the results are presented in Figure 5 with the adjusted odds ratio colour coded to indicate strength of association.

**Figure 5.**
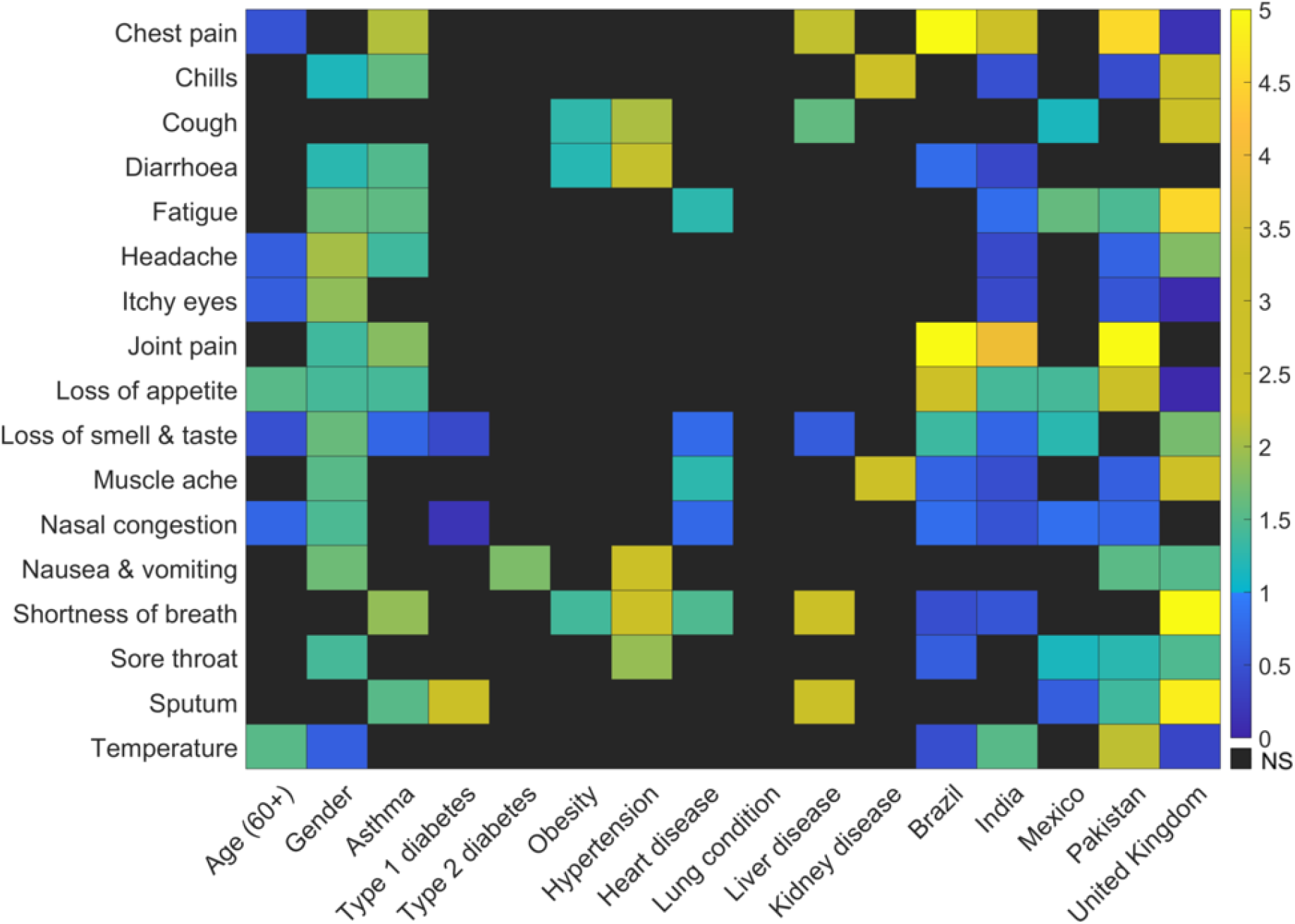
Association between symptoms, underlying chronic disease, countries, age and gender in tested positive responders. Each row represents the adjusted odds ratio of the different factors (age, gender, chronic disease and countries) in a multivariable model for the presence of the symptom corresponding to the row. Odds ratio which are statistically significant (p<0.05, 95% CI excludes 1) are coloured. Odds ratio which are not statistically significant are marked as NS.

After adjustment for other chronic diseases, gender, age and country of residence, COVID-19 positive responders with asthma or liver disease had the greatest odds of reporting chest pain and sputum production. COVID-19 positive responders with hypertension had the greatest odds of reporting shortness of breath, nausea and vomiting, sore throat, diarrhoea, or cough, compared with other chronic condition, and after adjustment for confounders.

COVID-19 responders from Brazil had the greatest odds of experiencing chest pain, joint pain and loss of appetite, compared with other countries and whilst controlling for age, gender and chronic diseases.

## DISCUSSION

This is the first study to look at symptoms among those who test positive for COVID-19 by geolocation and underlying chronic disease. We find that there are geographic and underlying disease symptom differences, and this understanding is crucial for clinical practice: to speed up diagnosis; enable more precise prediction of outcomes, and target treatment. Symptoms for COVID-19 positive patients varied based on underlying chronic disease and based on geographical location in both crude and adjusted logistic regression models.

### Previous work and how our results compare with others

The global impact of the COVID-19 pandemic has led to a rapid development and utilization of mobile health applications. To facilitate an agile response to the pandemic, self-reported survey responses on health, behaviour and demographics have been introduced in countries around the world to better understand symptom presentation. However, evidence suggests that their conclusions are highly variable. A recent Cochrane review found data on 84 signs and symptoms in 44 studies, but only 10 symptoms were reported by more than 10 studies. The top ten most often reported signs and symptoms were fever, cough, shortness of breath, sore throat, muscle soreness, diarrhoea, headache, fatigue, sputum production, and loss of smell or taste. (12) An early meta-analysis of epidemiological variation in COVID-19 inside and outside China studied patient characteristics including, gender, age, fatality rate, and symptoms of fever, cough, shortness of breath and diarrhoea in COVID-19 patients. They found that important symptom differences existed in patients in China compared to other countries and recommended that clinical symptoms of COVID-19 should not be generalized to fever, shortness of breath and cough only, but other symptoms such as diarrhoea are also shown to be prevalent in patients with COVID-19. (13) The use of symptom profiles to predict COVID-19 positive PCR has also had mixed results. A recent evaluation of the diagnostic accuracies of web-based COVID-19 symptom checkers found a variation in sensitivity and specificity of symptoms mappers meta-analysed. This was in part due to the wide variation in symptoms that could be added to individual symptom mappers.(1)

Responders from India comprised a high proportion of our study cohort and our study shows that amongst the Indian COVID-19 positive responders, fatigue, joint pain, muscle ache and headache were most commonly reported. A survey conducted in August 2020 in the Indian population aimed to assess knowledge and awareness on COVID-19.(14) Questions on awareness about coronavirus symptoms revealed that considerable numbers of respondents acknowledged fever, and persistent cough as frequent symptoms of COVID-19, but relatively few responders could list additional symptoms. This highlights a lack of knowledge on the most prevalent symptoms within that country.

A recent Brazilian study aimed to analyse the profile of COVID-19 symptoms and related aspects. Using data from 346,181 individuals who completed the *Brazilian National Household Sample Survey* in May 2020. (15) Eleven key symptoms were reported which were widely reported in the population: fever, cough, sore throat, difficulty breathing, headache, chest pain, nausea, stuffy or runny nose, fatigue, eye pain and loss of smell or taste. Female sex, brown skin colour, the North and Northeast regions of Brazil, and all three older age brackets showed stronger association with all the symptoms. Our results agree with the profile of symptoms reported in the Brazilian population, and we also observe age related increase in odds of symptoms, across all countries.

An observational study on 482,413 individuals who were tested for COVID-19 in Mexico found high incidence in working-age Mexican outpatients, the main symptoms among people tested were headache, arthralgia or myalgia, and sore throat. (16) The study identified a further 8 symptoms associated with COVID-19: abdominal pain, breathing difficulty, chills, conjunctivitis, constant dry cough, diarrhoea, fever, runny nose, or vomiting. Using clustering techniques 3 symptomatic profiles were suggested which grouped the 11 symptoms. These symptoms correspond to the symptoms recorded in our study. In our study Mexican responders with a COVID-19 positive result frequently reported fatigue, headache, itchy eyes and sort throat. In addition, our study found a that loss of smell and taste was reported more frequently in the Mexican COVID-19 positive responders than in responders from other countries.

Public health guidance in the UK advises that the triad of symptoms: new persistent cough; high temperature, loss of taste and smell. (17) In the UK a national symptom tracker app collects data from both asymptomatic and symptomatic individuals and tracks in real time how the disease progresses by recording self-reported health information on a daily basis, including symptoms, hospitalization, reverse-transcription PCR (RT-PCR) test outcomes, demographic information and pre-existing medical conditions. A recent report based on 2,618,862 individuals who used the app identified a combination of symptoms, in addition to the more stablished symptoms, including anosmia, fatigue, persistent cough and loss of appetite, that together might identify individuals with COVID-19. (18)

These results were further affirmed by the UK REACT (REal-time Assessment of Community Transmission). In addition to previously reported symptoms which were predictive of COVID-19 positive PCR the REACT programme reports that chills, headache, appetite loss and muscle aches should be added to the catalogue of COVID-19 symptoms. (19) The study also found that there was a variation in symptoms with age. While chills were linked with testing positive across all ages, headaches were reported in young people aged five to 17 and appetite loss was reported more in 18-54 year olds and those aged 55 and over. Cough was observed in two thirds of cases in a systematic review and the largest cohort studies, suggesting it is unreliable alone as a key diagnostic symptom. Our findings support these results, with chills, loss of smell and taste, fatigue, cough and loss of appetite being reported as prevalent symptoms in the UK responding population in our study. Muscle ache was more prevalent in the UK COVID-19 positive responders compared with other countries. In the REACT study muscle ache was mostly reported in people aged between 18 and 54. (17) Our study supports this finding, and this may reflect the older demographic of the UK population within our study.

### Strengths & limitations

One of the strengths of this study was the ability to look globally at symptoms with a specific breakdown by nationality, allowing geolocation and culture/behavioural aspects to be investigated. Symptom reports were conducted in local languages (e.g. Portugese, Hindi, etc) thus increasing accessibility, however translations may not match exactly within cultural contexts, e.g. “pain” in Brazil is “joint ache” in UK (but not stomach pain). Given the data for this analysis came from an Internet based survey, there will be differential access, however only a very low effort was needed to partake given the questionnaire was accessed via a simple website and not an app. Given the widespread use of smartphones globally, this should facilitate participation, however we acknowledge that those who are younger or in wealthier countries may be more likely to partake thus skewing the results, equally educational factors may have played a role and we do not have any socioeconomic or ethnicity information. Whilst we acknowledge that the data used are self-reported, we do not think this undermines the accuracy of underlying disease or symptom reporting. For those who report a COVID-19 positive test, we do not distinguish between type of tests and thus cannot account for differences in accuracy.

### Clinical and Public Health impact

Our information may be utilised in a clinical setting as an additional triage tool and for target testing, especially to better inform decisions in patient groups with preconditions, co- and multi-morbidities, but also in countries where no published symptom profiles have been reported. Symptom checkers are being widely used in response to the global COVID-19 pandemic. A recent study reported that web-based COVID-19 symptom checkers vary widely in their predictive capabilities, with some performing equivalently to random guessing while others show strength in sensitivity, specificity, or both. (1)

The results have wider public health implications beyond direct clinical care. The study highlights the importance of looking beyond the classic public health messaging on the classic symptoms of COVID-19. Our study corroborates findings from individual countries that a wide variety of symptoms are associated with COVID-19. Public health messaging in many countries is focussed on advice to seek a COVID-19 test if the triad of symptoms cough, fever and loss of taste/smell are present or variations thereof. The results indicate that amongst those patients with a COVID-19 positive test, and regardless of underlying chronic disease the most frequently reported symptoms did not include cough, fever and loss of taste/smell. Mismatches between the symptoms that populations were communicated to look out for and those they are actually presenting may mislead patients to assume an asymptomatic behavioural stance while they may be actually COVID-19 positive. If population testing around the world is triggered by symptom criteria that are inaccurate this could potentially bias any prevalence estimates, leading to an underrepresentation of COVID-19 positive cases, which will hamper measures to control and manage the pandemic.

## Data Availability

The Covid-10 Symptom Mapper data was provided to us by Your.MD free of charge and obligations with freedom to publish any results. The data is provided on request for free from Your.MD.

## Contributions

Study concepts & design: BK, GB, JKQ, CEC, AAF

Literature search: CEC

Data cleaning: BK, GB

Statistical Analysis: BK, GB

Data Analysis: BK, GB, JKQ, CEC, AAF

Manuscript draft: All authors

Figure preparation: BK, GB

Final manuscript review: All authors

## Acknowledgements

We thank Your.MD, Matteo Berlucchi and his team, for sharing their data with the public freely. We acknowledge funding to AAF (UKRI Turing AI Fellowship), CEC by a personal NIHR Career Development Fellowship (grant number NIHR-2016-090-015). JKQ has received grants from The Health Foundation, MRC, GSK, Bayer, BI, British Lung Foundation, IQVIA, Chiesi AZ, Insmed and Asthma UK. This work is supported by BREATHE - The Health Data Research Hub for Respiratory Health [MC_PC_19004]. BREATHE is funded through the UK Research and Innovation Industrial Strategy Challenge Fund and delivered through Health Data Research UK. Imperial College London is grateful for the support from the North West London NIHR Applied Research Collaboration. The funders had no role in study design or writing. All authors had full access to all the data in the study. The writing committee had sole responsibility for the decision to submit for publication.

**Disclaimer The views expressed in this publication are those of the authors and not necessarily those of the NIHR or the Department of Health and Social Care**.

## Supplementary Material

**Supplementary Figure S1.**
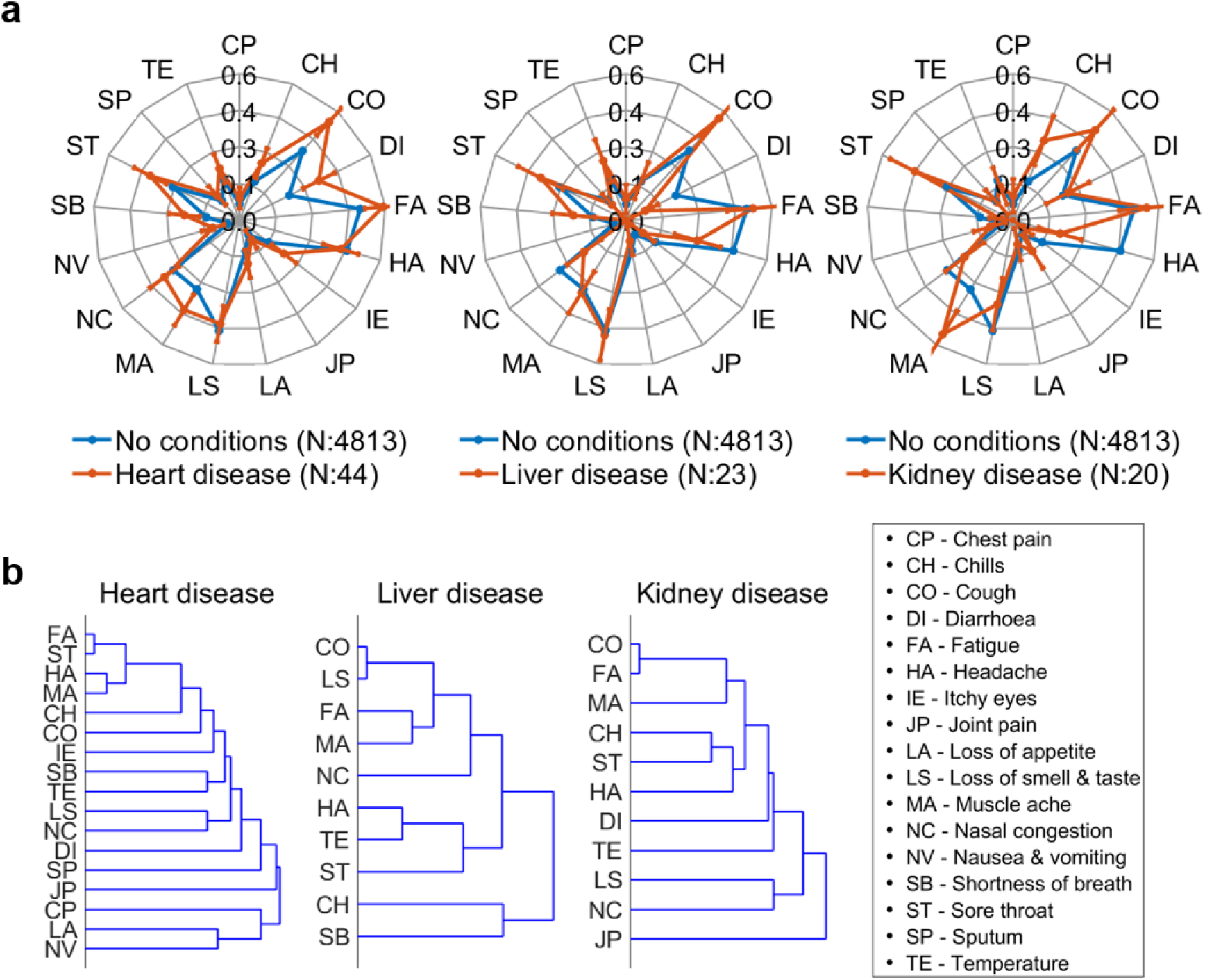
a. Symptom profile difference between no conditions group and each comorbidity group. Error bars shown are standard errors. b. Dendrograms of clustering of the symptoms for different comorbidity groups amongst COVID-19 positive responders

**Supplementary Table S1:** Survey questions asked at the YourMD symptom mapper webpage.

| Questions on Your.md website | Choose from options |
| --- | --- |
| First of all, it's important to find out if you need urgent attention. Are you experiencing any of the following? | Severe shortness of breath - more than 25 breaths per minute<br>Pain or pressure in your chest<br>Cold clammy or pale and mottled skin<br>Feeling confused (if that's new)<br>Coughing up blood<br>Blue lips or face<br>Sudden collapse or fainting<br>Difficulty waking up<br>Little or no urine output |
| What's the main reason you'd like to map your symptoms? Please choose the most relevant reason only. | I've tested positive for coronavirus and have symptoms<br>I tested negative but now have symptoms<br>I have symptoms but haven't been tested<br>I've recovered from coronavirus but have new symptoms<br>I'm just interested in seeing the symptoms comparison, I don't have symptoms<br>I am isolating but have no symptoms |
| How are you feeling? | I feel fine<br>I feel sick<br>I feel very sick |
| How many days have you had symptoms? | From 0 to 'more than 42' |
| Now for the important bit. Do you have any of these symptoms? [Multiple Answer Tick Boxes] | Cough<br>Fever<br>Fatigue/tiredness<br>Headache<br>Blocked nose<br>Diarrhoea<br>Loss of appetite<br>Chest pain /tightness<br>Joint pain /aches<br>Coughing up mucus<br>Shortness of breath<br>Muscular aches<br>Sore throat<br>Chills<br>Vomiting<br>Loss of taste and smell<br>Sneezing<br>Itchy eyes |
| We are keen to understand how COVID-19 is affecting people with long-standing health problems. Do you have any of these conditions?<br>[Multiple Answer Tick Boxes] | Long-standing lung condition<br>Asthma (managed with inhaler)<br>Long standing heart Disease<br>High Blood Pressure (hypertension)<br>Diabetes Type 1 (controlled by insulin)<br>Diabetes Type 2<br>Long-Standing Kidney Disease<br>Long Standing Liver Disease<br>Obesity |
| Take a look at these descriptions of mild, moderate and severe COVID-19. At the end, tell us which one best describes how you're feeling now. | Mild COVID-19<br>How it feels: You'll have flu-like symptoms, which can include cough, mild fever, headache, muscle pain and tiredness. You won't be short of breath in general. You shouldn't have a high fever. Keep yourself hydrated and rest. More info...<br>Moderate COVID-19<br>How it feels: You're more breathless and feel your heart beat faster when you move around. The cough is more noticeable. You'll be coughing many times an hour and it's probably making you tired. You may have symptoms of a fever like shivering and chills. More info...<br>Severe COVID-19<br>How it feels: You're going to feel very breathless (even when you're resting), and might find it hard to finish sentences. Along with chest, stomach or back pain when you breathe, these can be signs you've developed pneumonia. You will not be eating and drinking normally. Other signs include a high fever. More info...<br>Warning: If you're feeling increasingly breathless, can't manage basic things like showering and eating, or if you can't speak in whole sentences without taking extra breaths, call your local health service right away.<br>*Read Dr Mary Lowth's authoritative article on the stages of coronavirus on Healthily |
| After reading this, would you say your symptoms are | Mild; Moderate, Severe, None of the above |
| Town/City and Country |  |
| Age |  |
| Sex at birth | Male; Female |
| Are you pregnant? |  |
| Are you a healthcare worker? |  |
| Are you a care home worker? |  |

**Supplementary Table S2:** Symptoms and comorbidities reported by tested positive responders from top 5 countries

|  | SARS-COVID-19 status |  |  |  |
| --- | --- | --- | --- | --- |
|  | All Responders | Tested Positive | Untested Symptomatic | Tested Negative |
| **Brazil** |  |  |  |  |
| Answered questions on symptoms | 7257 | 1366 | 5089 | 802 |
| Chest pain (n/%) | 1095(15.1) | 230(16.8) | 708(13.9) | 157(19.6) |
| Chills (n/%) | 1587(21.9) | 255(18.7) | 1144(22.5) | 188(23.4) |
| Cough (n/%) | 2856(39.4) | 532(38.9) | 2016(39.6) | 308(38.4) |
| Diarrhoea (n/%) | 1548(21.3) | 327(23.9) | 1038(20.4) | 183(22.8) |
| Fatigue (n/%) | 3096(42.7) | 664(48.6) | 2074(40.8) | 358(44.6) |
| Headache (n/%) | 3889(53.6) | 691(50.6) | 2748(54.0) | 450(56.1) |
| Itchy eyes (n/%) | 1558(21.5) | 259(19.0) | 1109(21.8) | 190(23.7) |
| Joint pain (n/%) | 1491(20.5) | 261(19.1) | 1052(20.7) | 178(22.2) |
| Loss of Appetite (n/%) | 1461(20.1) | 331(24.2) | 953(18.7) | 177(22.1) |
| Loss of smell and taste(n/%) | 2453(33.8) | 695(50.9) | 1510(29.7) | 248(30.9) |
| Muscle Ache (n/%) | 2315(31.9) | 401(29.4) | 1648(32.4) | 266(33.2) |
| Nasal Congestion (n/%) | 3203(44.1) | 458(33.5) | 2390(47.0) | 355(44.3) |
| Nausea and vomiting (n/%) | 300(4.1) | 65(4.8) | 199(3.9) | 36(4.5) |
| Shortness of breath (n/%) | 582(8.0) | 114(8.3) | 385(7.6) | 83(10.3) |
| Sore throat (n/%) | 2557(35.2) | 313(22.9) | 1961(38.5) | 283(35.3) |
| Sputum (n/%) | 895(12.3) | 135(9.9) | 671(13.2) | 89(11.1) |
| Temperature(n/%) | 965(13.3) | 122(8.9) | 740(14.5) | 103(12.8) |
| Answered questions on chronic conditions | 7257 | 1366 | 5089 | 802 |
| Reported one or more chronic conditions | 3026(41.7) | 556(40.7) | 2121(41.7) | 349(43.5) |
| Asthma (%) | 425(5.9) | 74(5.4) | 302(5.9) | 49(6.1) |
| Diabetes type I (%) | 54(0.7) | 13(1.0) | 30(0.6) | 11(1.4) |
| Diabetes type II (%) | 262(3.6) | 56(4.1) | 170(3.3) | 36(4.5) |
| Heart disease (%) | 120(1.7) | 24(1.8) | 76(1.5) | 20(2.5) |
| Hypertension (%) | 1460(20.1) | 281(20.6) | 993(19.5) | 186(23.2) |
| Kidney disease (%) | 41(0.6) | 5(0.4) | 28(0.6) | 8(1.0) |
| Liver disease (%) | 200(2.8) | 23(1.7) | 162(3.2) | 15(1.9) |
| Lung condition (%) | 65(0.9) | 3(0.2) | 58(1.1) | 4(0.5) |
| Obesity (%) | 1395(19.2) | 248(18.2) | 982(19.3) | 165(20.6) |
| **India** |  |  |  |  |
| Answered questions on symptoms | 8967 | 714 | 7552 | 701 |
| Chest pain (n/%) | 494(5.5) | 60(8.4) | 383(5.1) | 51(7.3) |
| Chills (n/%) | 1015(11.3) | 72(10.1) | 883(11.7) | 60(8.6) |
| Cough (n/%) | 3433(38.3) | 266(37.3) | 2950(39.1) | 217(31.0) |
| Diarrhoea (n/%) | 910(10.1) | 90(12.6) | 761(10.1) | 59(8.4) |
| Fatigue (n/%) | 3577(39.9) | 280(39.2) | 3073(40.7) | 224(32.0) |
| Headache (n/%) | 2567(28.6) | 163(22.8) | 2251(29.8) | 153(21.8) |
| Itchy eyes (n/%) | 504(5.6) | 50(7.0) | 414(5.5) | 40(5.7) |
| Joint pain (n/%) | 512(5.7) | 57(8.0) | 415(5.5) | 40(5.7) |
| Loss of Appetite (n/%) | 525(5.9) | 83(11.6) | 397(5.3) | 45(6.4) |
| Loss of smell and taste(n/%) | 1801(20.1) | 228(31.9) | 1478(19.6) | 95(13.6) |
| Muscle Ache (n/%) | 2199(24.5) | 139(19.5) | 1909(25.3) | 151(21.5) |
| Nasal Congestion (n/%) | 2413(26.9) | 151(21.1) | 2081(27.6) | 181(25.8) |
| Nausea and vomiting (n/%) | 363(4.0) | 39(5.5) | 299(4.0) | 25(3.6) |
| Shortness of breath (n/%) | 1401(15.6) | 62(8.7) | 1225(16.2) | 114(16.3) |
| Sore throat (n/%) | 3669(40.9) | 181(25.4) | 3250(43.0) | 238(34.0) |
| Sputum (n/%) | 1714(19.1) | 92(12.9) | 1506(19.9) | 116(16.5) |
| Temperature(n/%) | 2017(22.5) | 206(28.9) | 1674(22.2) | 137(19.5) |
| Answered questions on chronic conditions | 8967 | 714 | 7552 | 701 |
| Reported one or more chronic conditions | 2903(32.4) | 281(39.4) | 2331(30.9) | 291(41.5) |
| Asthma (%) | 318(3.5) | 23(3.2) | 261(3.5) | 34(4.9) |
| Diabetes type I (%) | 116(1.3) | 20(2.8) | 87(1.2) | 9(1.3) |
| Diabetes type II (%) | 454(5.1) | 82(11.5) | 325(4.3) | 47(6.7) |
| Heart disease (%) | 91(1.0) | 18(2.5) | 63(0.8) | 10(1.4) |
| Hypertension (%) | 906(10.1) | 94(13.2) | 737(9.8) | 75(10.7) |
| Kidney disease (%) | 86(1.0) | 6(0.8) | 66(0.9) | 14(2.0) |
| Liver disease (%) | 179(2.0) | 32(4.5) | 122(1.6) | 25(3.6) |
| Lung condition (%) | 70(0.8) | 13(1.8) | 47(0.6) | 10(1.4) |
| Obesity (%) | 1316(14.7) | 99(13.9) | 1084(14.4) | 133(19.0) |
| **Mexico** |  |  |  |  |
| Answered questions on symptoms | 20616 | 2199 | 17432 | 985 |
| Chest pain (n/%) | 444(2.2) | 76(3.5) | 343(2.0) | 25(2.5) |
| Chills (n/%) | 4182(20.3) | 385(17.5) | 3593(20.6) | 204(20.7) |
| Cough (n/%) | 8053(39.1) | 929(42.2) | 6757(38.8) | 367(37.3) |
| Diarrhoea (n/%) | 4724(22.9) | 581(26.4) | 3920(22.5) | 223(22.6) |
| Fatigue (n/%) | 10808(52.4) | 1270(57.8) | 9027(51.8) | 511(51.9) |
| Headache (n/%) | 10877(52.8) | 1073(48.8) | 9311(53.4) | 493(50.1) |
| Itchy eyes (n/%) | 3484(16.9) | 393(17.9) | 2917(16.7) | 174(17.7) |
| Joint pain (n/%) | 413(2.0) | 59(2.7) | 326(1.9) | 28(2.8) |
| Loss of Appetite (n/%) | 1664(8.1) | 272(12.4) | 1288(7.4) | 104(10.6) |
| Loss of smell and taste(n/%) | 7786(37.8) | 1038(47.2) | 6434(36.9) | 314(31.9) |
| Muscle Ache (n/%) | 8158(39.6) | 831(37.8) | 6957(39.9) | 370(37.6) |
| Nasal Congestion (n/%) | 8113(39.4) | 714(32.5) | 7037(40.4) | 362(36.8) |
| Nausea and vomiting (n/%) | 980(4.8) | 107(4.9) | 838(4.8) | 35(3.6) |
| Shortness of breath (n/%) | 2601(12.6) | 328(14.9) | 2165(12.4) | 108(11.0) |
| Sore throat (n/%) | 9055(43.9) | 721(32.8) | 7910(45.4) | 424(43.0) |
| Sputum (n/%) | 1607(7.8) | 156(7.1) | 1376(7.9) | 75(7.6) |
| Temperature(n/%) | 4774(23.2) | 441(20.1) | 4137(23.7) | 196(19.9) |
| Answered questions on chronic conditions | 20616 | 2199 | 17432 | 985 |
| Reported one or more chronic conditions | 8256(40.0) | 913(41.5) | 6904(39.6) | 439(44.6) |
| Asthma (%) | 763(3.7) | 87(4.0) | 633(3.6) | 43(4.4) |
| Diabetes type I (%) | 260(1.3) | 32(1.5) | 213(1.2) | 15(1.5) |
| Diabetes type II (%) | 968(4.7) | 132(6.0) | 778(4.5) | 58(5.9) |
| Heart disease (%) | 181(0.9) | 12(0.5) | 149(0.9) | 20(2.0) |
| Hypertension (%) | 2306(11.2) | 266(12.1) | 1895(10.9) | 145(14.7) |
| Kidney disease (%) | 72(0.3) | 6(0.3) | 62(0.4) | 4(0.4) |
| Liver disease (%) | 182(0.9) | 28(1.3) | 142(0.8) | 12(1.2) |
| Lung condition (%) | 103(0.5) | 9(0.4) | 87(0.5) | 7(0.7) |
| Obesity (%) | 5468(26.5) | 595(27.1) | 4620(26.5) | 253(25.7) |
| **Pakistan** |  |  |  |  |
| Answered questions on symptoms | 7451 | 753 | 6286 | 412 |
| Chest pain (n/%) | 718(9.6) | 96(12.7) | 578(9.2) | 44(10.7) |
| Chills (n/%) | 885(11.9) | 75(10.0) | 753(12.0) | 57(13.8) |
| Cough (n/%) | 3259(43.7) | 308(40.9) | 2770(44.1) | 181(43.9) |
| Diarrhoea (n/%) | 1086(14.6) | 172(22.8) | 864(13.7) | 50(12.1) |
| Fatigue (n/%) | 4236(56.9) | 398(52.9) | 3596(57.2) | 242(58.7) |
| Headache (n/%) | 2811(37.7) | 253(33.6) | 2398(38.1) | 160(38.8) |
| Itchy eyes (n/%) | 793(10.6) | 65(8.6) | 683(10.9) | 45(10.9) |
| Joint pain (n/%) | 1131(15.2) | 119(15.8) | 952(15.1) | 60(14.6) |
| Loss of Appetite (n/%) | 826(11.1) | 125(16.6) | 649(10.3) | 52(12.6) |
| Loss of smell and taste(n/%) | 2325(31.2) | 311(41.3) | 1907(30.3) | 107(26.0) |
| Muscle Ache (n/%) | 2161(29.0) | 189(25.1) | 1850(29.4) | 122(29.6) |
| Nasal Congestion (n/%) | 2305(30.9) | 207(27.5) | 1976(31.4) | 122(29.6) |
| Nausea and vomiting (n/%) | 412(5.5) | 57(7.6) | 332(5.3) | 23(5.6) |
| Shortness of breath (n/%) | 1028(13.8) | 114(15.1) | 859(13.7) | 55(13.3) |
| Sore throat (n/%) | 3103(41.6) | 247(32.8) | 2679(42.6) | 177(43.0) |
| Sputum (n/%) | 1430(19.2) | 118(15.7) | 1225(19.5) | 87(21.1) |
| Temperature(n/%) | 2736(36.7) | 270(35.9) | 2327(37.0) | 139(33.7) |
| Answered questions on chronic conditions | 7451 | 753 | 6286 | 412 |
| Reported one or more chronic preconditions | 2594(34.8) | 302(40.1) | 2115(33.6) | 177(43.0) |
| Asthma (%) | 342(4.6) | 36(4.8) | 284(4.5) | 22(5.3) |
| Diabetes type I (%) | 128(1.7) | 27(3.6) | 95(1.5) | 6(1.5) |
| Diabetes type II (%) | 355(4.8) | 48(6.4) | 284(4.5) | 23(5.6) |
| Heart disease (%) | 93(1.2) | 15(2.0) | 72(1.1) | 6(1.5) |
| Hypertension (%) | 765(10.3) | 103(13.7) | 605(9.6) | 57(13.8) |
| Kidney disease (%) | 66(0.9) | 11(1.5) | 51(0.8) | 4(1.0) |
| Liver disease (%) | 194(2.6) | 29(3.9) | 151(2.4) | 14(3.4) |
| Lung condition (%) | 82(1.1) | 8(1.1) | 69(1.1) | 5(1.2) |
| Obesity (%) | 1184(15.9) | 117(15.5) | 993(15.8) | 74(18.0) |
| **United Kingdom** |  |  |  |  |
| Answered questions on symptoms | 2977 | 420 | 2379 | 178 |
| Chest pain (n/%) | 27(0.9) | 3(0.7) | 23(1.0) | 1(0.6) |
| Chills (n/%) | 1152(38.7) | 159(37.9) | 928(39.0) | 65(36.5) |
| Cough (n/%) | 1772(59.5) | 257(61.2) | 1405(59.1) | 110(61.8) |
| Diarrhoea (n/%) | 787(26.4) | 100(23.8) | 640(26.9) | 47(26.4) |
| Fatigue (n/%) | 2334(78.4) | 338(80.5) | 1862(78.3) | 134(75.3) |
| Headache (n/%) | 1826(61.3) | 270(64.3) | 1448(60.9) | 108(60.7) |
| Itchy eyes (n/%) | 44(1.5) | 7(1.7) | 33(1.4) | 4(2.2) |
| Joint pain (n/%) | 43(1.4) | 7(1.7) | 35(1.5) | 1(0.6) |
| Loss of Appetite (n/%) | 32(1.1) | 2(0.5) | 29(1.2) | 1(0.6) |
| Loss of smell and taste(n/%) | 982(33.0) | 243(57.9) | 689(29.0) | 50(28.1) |
| Muscle Ache (n/%) | 1835(61.6) | 276(65.7) | 1448(60.9) | 111(62.4) |
| Nasal Congestion (n/%) | 1163(39.1) | 166(39.5) | 935(39.3) | 62(34.8) |
| Nausea and vomiting (n/%) | 304(10.2) | 39(9.3) | 241(10.1) | 24(13.5) |
| Shortness of breath (n/%) | 1370(46.0) | 223(53.1) | 1068(44.9) | 79(44.4) |
| Sore throat (n/%) | 1535(51.6) | 174(41.4) | 1277(53.7) | 84(47.2) |
| Sputum (n/%) | 1180(39.6) | 157(37.4) | 962(40.4) | 61(34.3) |
| Temperature(n/%) | 263(8.8) | 31(7.4) | 217(9.1) | 15(8.4) |
| Answered questions on chronic conditions | 2977 | 420 | 2379 | 178 |
| Reported one or more chronic conditions | 1020(34.3) | 152(36.2) | 800(33.6) | 68(38.2) |
| Asthma (%) | 351(11.8) | 54(12.9) | 272(11.4) | 25(14.0) |
| Diabetes type I (%) | 18(0.6) | 3(0.7) | 14(0.6) | 1(0.6) |
| Diabetes type II (%) | 125(4.2) | 19(4.5) | 98(4.1) | 8(4.5) |
| Heart disease (%) | 49(1.6) | 4(1.0) | 40(1.7) | 5(2.8) |
| Hypertension (%) | 372(12.5) | 60(14.3) | 291(12.2) | 21(11.8) |
| Kidney disease (%) | 15(0.5) | 1(0.2) | 11(0.5) | 3(1.7) |
| Liver disease (%) | 76(2.6) | 6(1.4) | 64(2.7) | 6(3.4) |
| Lung condition (%) | 20(0.7) | 4(1.0) | 14(0.6) | 2(1.1) |
| Obesity (%) | 383(12.9) | 60(14.3) | 295(12.4) | 28(15.7) |

**Supplementary Table S3:**
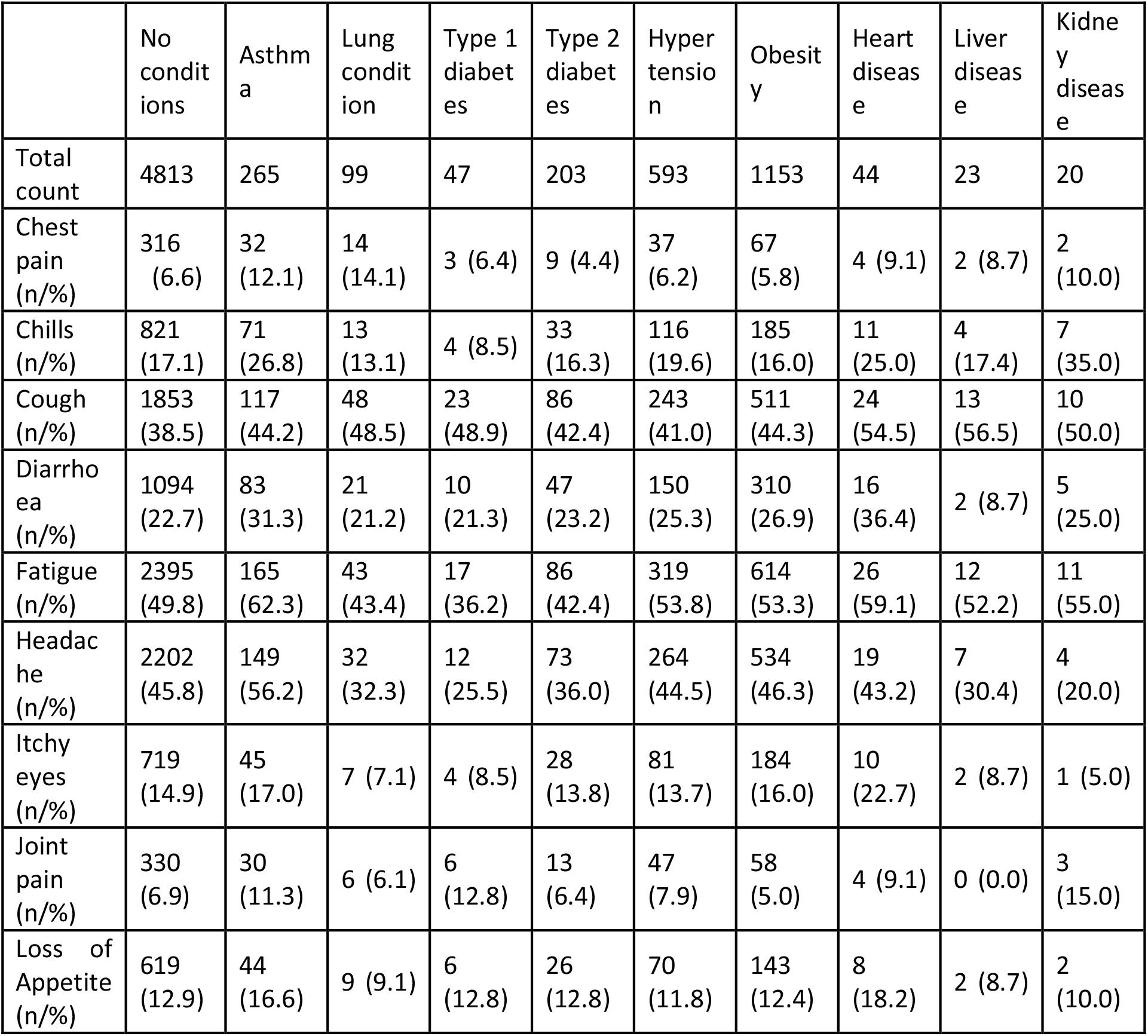
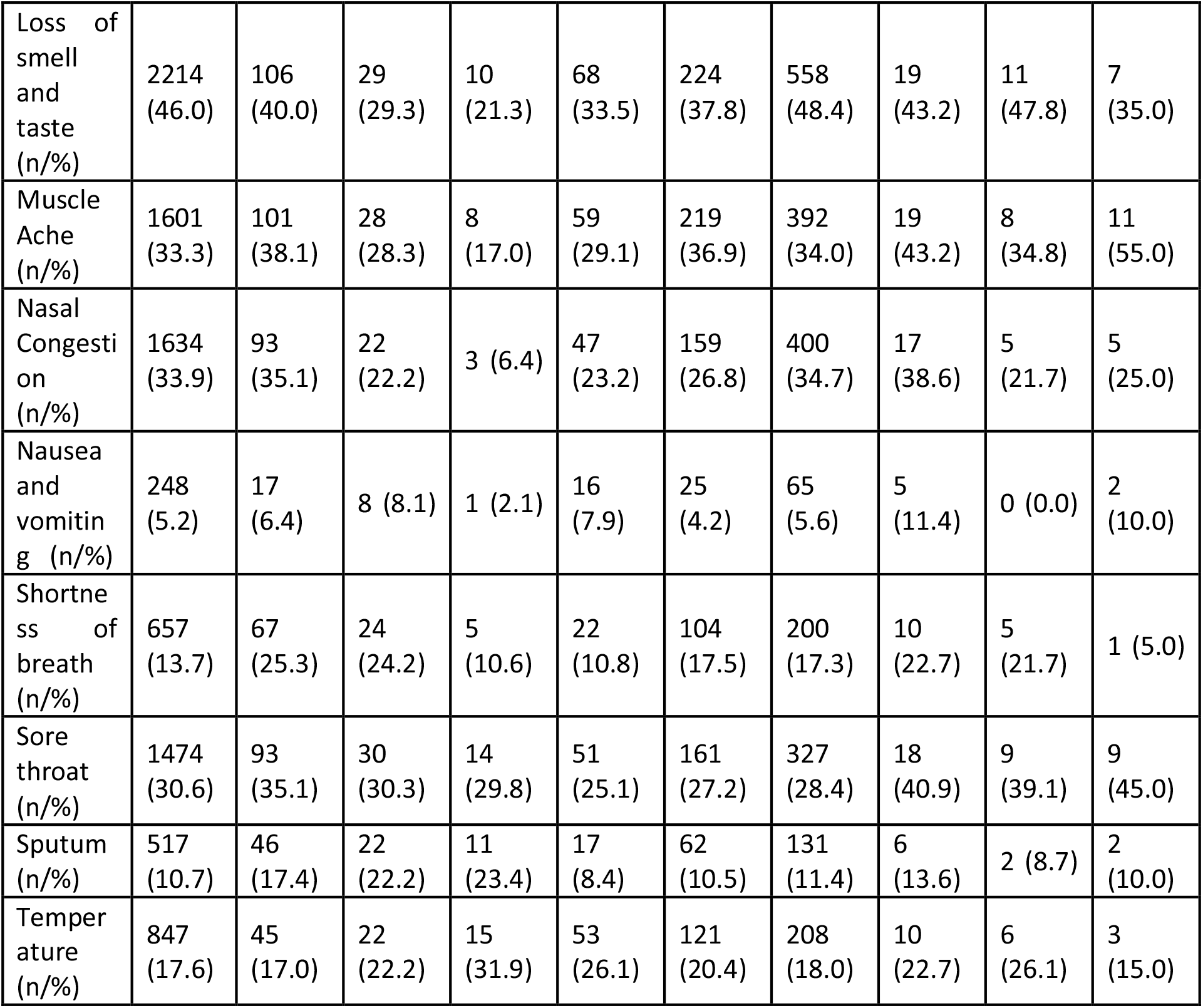
Symptoms reported by tested positive respondents with underlying chronic preconditions.

|  | No conditions | Asthma | Lung condition | Type 1 diabetes | Type 2 diabetes | Hypertension | Obesity | Heart disease | Liver disease | Kidney disease |
| --- | --- | --- | --- | --- | --- | --- | --- | --- | --- | --- |
| Total count | 4813 | 265 | 99 | 47 | 203 | 593 | 1153 | 44 | 23 | 20 |
| Chest pain (n/%) | 316 (6.6) | 32 (12.1) | 14 (14.1) | 3 (6.4) | 9 (4.4) | 37 (6.2) | 67 (5.8) | 4 (9.1) | 2 (8.7) | 2 (10.0) |
| Chills (n/%) | 821 (17.1) | 71 (26.8) | 13 (13.1) | 4 (8.5) | 33 (16.3) | 116 (19.6) | 185 (16.0) | 11 (25.0) | 4 (17.4) | 7 (35.0) |
| Cough (n/%) | 1853 (38.5) | 117 (44.2) | 48 (48.5) | 23 (48.9) | 86 (42.4) | 243 (41.0) | 511 (44.3) | 24 (54.5) | 13 (56.5) | 10 (50.0) |
| Diarrhoea (n/%) | 1094 (22.7) | 83 (31.3) | 21 (21.2) | 10 (21.3) | 47 (23.2) | 150 (25.3) | 310 (26.9) | 16 (36.4) | 2 (8.7) | 5 (25.0) |
| Fatigue (n/%) | 2395 (49.8) | 165 (62.3) | 43 (43.4) | 17 (36.2) | 86 (42.4) | 319 (53.8) | 614 (53.3) | 26 (59.1) | 12 (52.2) | 11 (55.0) |
| Headache (n/%) | 2202 (45.8) | 149 (56.2) | 32 (32.3) | 12 (25.5) | 73 (36.0) | 264 (44.5) | 534 (46.3) | 19 (43.2) | 7 (30.4) | 4 (20.0) |
| Itchy eyes (n/%) | 719 (14.9) | 45 (17.0) | 7 (7.1) | 4 (8.5) | 28 (13.8) | 81 (13.7) | 184 (16.0) | 10 (22.7) | 2 (8.7) | 1 (5.0) |
| Joint pain (n/%) | 330 (6.9) | 30 (11.3) | 6 (6.1) | 6 (12.8) | 13 (6.4) | 47 (7.9) | 58 (5.0) | 4 (9.1) | 0 (0.0) | 3 (15.0) |
| Loss of Appetite (n/%) | 619 (12.9) | 44 (16.6) | 9 (9.1) | 6 (12.8) | 26 (12.8) | 70 (11.8) | 143 (12.4) | 8 (18.2) | 2 (8.7) | 2 (10.0) |
| Loss of smell and taste (n/%) | 2214 (46.0) | 106 (40.0) | 29 (29.3) | 10 (21.3) | 68 (33.5) | 224 (37.8) | 558 (48.4) | 19 (43.2) | 11 (47.8) | 7 (35.0) |
| Muscle Ache (n/%) | 1601 (33.3) | 101 (38.1) | 28 (28.3) | 8 (17.0) | 59 (29.1) | 219 (36.9) | 392 (34.0) | 19 (43.2) | 8 (34.8) | 11 (55.0) |
| Nasal Congestion (n/%) | 1634 (33.9) | 93 (35.1) | 22 (22.2) | 3 (6.4) | 47 (23.2) | 159 (26.8) | 400 (34.7) | 17 (38.6) | 5 (21.7) | 5 (25.0) |
| Nausea and vomiting (n/%) | 248 (5.2) | 17 (6.4) | 8 (8.1) | 1 (2.1) | 16 (7.9) | 25 (4.2) | 65 (5.6) | 5 (11.4) | 0 (0.0) | 2 (10.0) |
| Shortness of breath (n/%) | 657 (13.7) | 67 (25.3) | 24 (24.2) | 5 (10.6) | 22 (10.8) | 104 (17.5) | 200 (17.3) | 10 (22.7) | 5 (21.7) | 1 (5.0) |
| Sore throat (n/%) | 1474 (30.6) | 93 (35.1) | 30 (30.3) | 14 (29.8) | 51 (25.1) | 161 (27.2) | 327 (28.4) | 18 (40.9) | 9 (39.1) | 9 (45.0) |
| Sputum (n/%) | 517 (10.7) | 46 (17.4) | 22 (22.2) | 11 (23.4) | 17 (8.4) | 62 (10.5) | 131 (11.4) | 6 (13.6) | 2 (8.7) | 2 (10.0) |
| Temperature (n/%) | 847 (17.6) | 45 (17.0) | 22 (22.2) | 15 (31.9) | 53 (26.1) | 121 (20.4) | 208 (18.0) | 10 (22.7) | 6 (26.1) | 3 (15.0) |

## Notes

### Competing Interest Statement

Dr. Quint reports grants from MRC, grants from GSK, grants and personal fees from AZ, grants and personal fees from BI, grants and personal fees from Chiesi, grants from The Health Foundation, grants from Bayer, grants from Asthma UK, outside the submitted work;.

### Author Declarations

The Covid-10 Symptom Mapper data was provided to us by the company Your.MD free of charge and obligations with freedom to publish any results. The data is provided on request for free from Your.MD. On the Your.MD website all participants provided informed consent at the start of the online questionnaire for their data to be used for research purposes, and had to agree to the corresponding Your.MD privacy and data usage policies.

